# A REAl-life study on short-term Dual Antiplatelet treatment in Patients with ischemic stroke or Transient ischemic attack (READAPT)

**DOI:** 10.1101/2024.01.08.24301025

**Authors:** Eleonora De Matteis, Raffaele Ornello, Federico De Santis, Matteo Foschi, Michele Romoli, Tiziana Tassinari, Valentina Saia, Silvia Cenciarelli, Chiara Bedetti, Chiara Padiglioni, Bruno Censori, Valentina Puglisi, Luisa Vinciguerra, Maria Guarino, Valentina Barone, Marialuisa Zedde, Ilaria Grisendi, Marina Diomedi, Maria Rosaria Bagnato, Marco Petruzzellis, Domenico Maria Mezzapesa, Pietro Di Viesti, Vincenzo Inchingolo, Manuel Cappellari, Mara Zenorini, Paolo Candelaresi, Vincenzo Andreone, Giuseppe Rinaldi, Alessandra Bavaro, Anna Cavallini, Stefan Morau, Pietro Querzani, Valeria Terruso, Marina Mannino, Alessandro Pezzini, Giovanni Frisullo, Francesco Muscia, Maurizio Paciaroni, Maria Giulia Mosconi, Andrea Zini, Ruggiero Leone, Carmela Palmieri, Letizia Maria Cupini, Michela Marcon, Rossana Tassi, Enzo Sanzaro, Cristina Paci, Giovanna Viticchi, Daniele Orsucci, Anne Falcou, Susanna Diamanti, Roberto Tarletti, Patrizia Nencini, Eugenia Rota, Federica Nicoletta Sepe, Delfina Ferrandi, Luigi Caputi, Gino Volpi, Salvatore La Spada, Mario Beccia, Claudia Rinaldi, Vincenzo Mastrangelo, Francesco Di Blasio, Paolo Invernizzi, Giuseppe Pelliccioni, Maria Vittoria De Angelis, Laura Bonanni, Giampietro Ruzza, Emanuele Alessandro Caggia, Monia Russo, Agnese Tonon, Maria Cristina Acciarri, Sabrina Anticoli, Cinzia Roberti, Giovanni Manobianca, Gaspare Scaglione, Francesca Pistoia, Alberto Fortini, Antonella De Boni, Alessandra Sanna, Alberto Chiti, Leonardo Barbarini, Marcella Caggiula, Maela Masato, Massimo Del Sette, Francesco Passarelli, Maria Roberta Bongioanni, Danilo Toni, Stefano Ricci, Simona Sacco, the READAPT Study Group

## Abstract

**Background and purpose:** Clinical trials proved the efficacy of short-term dual antiplatelet therapy (DAPT) in secondary prevention of minor ischemic stroke or high-risk transient ischemic attack (TIA). In clinical practice, the use of short-term DAPT is broader than in clinical trials and procedures of clinical trials (i.e. loading dose, timely treatment start) are not always followed. We aimed at evaluating effectiveness and safety of short-term DAPT in real-world.

**Methods:** READAPT (A REAl-life study on short-term Dual Antiplatelet treatment in Patients with ischemic stroke or Transient ischemic attack) (NCT05476081) was an observational multicenter real-world study with a 90-day follow-up. The enrollment closed on 28^th^ February 2023. We included patients aged 18+ receiving DAPT after minor ischemic stroke or high-risk TIA but no stringent exclusion criteria were applied regarding stroke severity and ABCD^2^ score. Primary effectiveness outcome was stroke (ischemic or hemorrhagic) or death due to vascular causes, primary safety outcome was moderate-to-severe bleeding. Key secondary outcomes were the type of ischemic and hemorrhagic events, disability, cause of death, and compliance to treatment.

**Results:** We included 1920 patients; the majority started DAPT after an ischemic stroke (69.9%) rather than a TIA. Primary effectiveness outcome occurred in 3.9% and primary safety outcome in 0.6% of cases. In total, 3.3% cerebrovascular ischemic recurrences occurred, 0.2% intracerebral hemorrhages, and 2.7% bleedings; 0.2% of patients died due to vascular causes. We compared different subgroups of patients and we found significant differences among patients with NIHSS score ≤5 and those without acute lesions at neuroimaging who had higher primary effectiveness outcomes than their counterparts. Additionally, DAPT start after 24 hours from symptom onset was associated with a lower likelihood of bleeding.

**Conclusions:** Short-term DAPT after ischemic stroke or TIA is effective and safe in real-world where a broader population receives the treatment and procedures may differ from clinical trials.

## 1. Introduction

Minor ischemic strokes and high-risk transient ischemic attacks (TIA) account for one half of all cerebrovascular ischemic events^1,2^ and are associated with a 4% risk of ischemic recurrences at 90 days^3-5^. The cornerstone of secondary prevention of minor stroke and high-risk TIA is short-term dual antiplatelet therapy (DAPT), which has proven superior to single antiplatelet therapy (SAPT) in reducing the risk of cerebrovascular ischemic recurrences when at its maximum^6-8^. Three randomized controlled trials (RCTs) showed that the benefits of DAPT - started within 12-24 hours from symptom onset - offset slightly increased hemorrhagic risks. Specifically, CHANCE and POINT assessed the efficacy of 21- and 90-day clopidogrel/aspirin DAPT in patients with NIHSS score ≤3 and ABCD^2^ score ≥4^6,7^, while THALES assessed the efficacy of 30-day ticagrelor/aspirin DAPT in patients with NIHSS score ≤5 and ABCD^2^ score ≥6 or symptomatic intra/extracranial arterial stenosis^8^. Recently, INSPIRES confirmed the efficacy and safety of 21-day clopidogrel/aspirin DAPT after an ischemic event of presumed atherosclerosis origin in patients with NIHSS score ≤5 and ABCD^2^ score ≥4 who started the treatment within 72 hours from symptom onset^9^.

Despite real-world cohort studies showed that DAPT use has increased since the publication of the RCTs^10^, around half of the eligible patients do not receive the treatment yet^11,12^ and there are disparities in treatment prescription – female gender, ethnicity, and cause of the index event are among the factors associated with lack of DAPT prescription^10^. Conversely, physicians occasionally prescribe DAPT to patients with non-minor stroke^11,13^ or who do not follow RCTs procedure^13^. A previous analysis of A REAl-life study on short-term Dual Antiplatelet treatment in patients with ischemic stroke or Transient ischemic attack (READAPT) study showed that only 8% of patients treated with DAPT in Italian stroke centers would have met RCTs inclusion criteria and followed their procedures^13^. Benefit/risk profile of DAPT in these patients had not been assessed yet. READAPT aimed at evaluating effectiveness and safety of DAPT in a multicenter real-world setting and in subgroups of patients who were excluded from RCTs (i.e. patients undergoing revascularization procedures, with more severe ischemic stroke, or lower risk TIA) or who did not follow RCTs procedures as per time to DAPT start, and antiplatelet loading dose.

## 2. Material and Methods

Study methodology has been previously reported^13^ and is briefly summarized in the following sections. READAPT (NCT05476081) is an observational prospective multicenter real-world study, which included a baseline observation (i.e. index event) and a 90-day follow-up. University of L’Aquila coordinated the study and Italian Stroke Association-Associazione Italiana Ictus (ISA-AII) endorsed it. The study adhered to the Strengthening the Reporting of Observational Studies in Epidemiology (STROBE) guidelines^14^. The Internal review Board of the University of L’Aquila approved the study with the number 03/2021 in February 2021 and ethic committees of each participating center subsequently cleared it. All included patients or their proxies signed an informed consent. Since February 2021, 64 Italian centers (Table S3) have enrolled all consecutive patients treated with DAPT. The enrolment closed on 28^th^ February 2023.

Patients were included shortly after the index event (i.e. baseline) and started a 90-day follow-up. At the end of follow-up period, local investigators performed a remote or face-to-face end of study visit.

### 2.1 Inclusion and exclusion criteria

Briefly, we included patients either hospitalized or not with non-cardioembolic minor ischemic stroke or high-risk TIA, older than 18 years, and receiving short-term DAPT. We referred to the World Health Organization (WHO) time-based criterion to distinguish ischemic strokes from TIAs. We did not set strict NIHSS and ABCD^2^ cut-off scores to exclude patients, but we recommended local investigators to adhere to guidelines^15-18^.

We excluded patients either receiving DAPT after endovascular stenting procedures, already randomized to interventional RCTs on stroke prevention, or who could have not ensured compliance to study procedures or data reliability.

### 2.2 Study procedures and data collection

Physicians chose DAPT regimen – loading dose and treatment duration – and the antiplatelet to be continued after DAPT discontinuation according to the best clinical practice. We encouraged a 21-30 day-course of DAPT, which could be extended up to 90 days upon RCTs evidence and guidelines^15-18^. We defined DAPT loading dose as aspirin 300 mg and/or clopidogrel 300-600 mg or ticagrelor 180 mg^6-8^. The loading dose was administered on day 1 of DAPT and followed by aspirin 100 mg/daily together with clopidogrel 75 mg/daily or ticagrelor 90 mg twice daily for the remaining DAPT period.

The baseline case-report-form collected patients’ demographics, vascular risk factors, pre-event treatment, NIHSS score, ABCD^2^ score, pre- and post-event modified Rankin Scale (mRS), time to DAPT start, treatment regimen, concurrent treatments, neuroimaging characteristics, acute treatment (i.e. thrombolysis, thrombectomy, endarterectomy), and cause of the index event according to the Trial of Org 10172 in acute stroke treatment (TOAST) classification^19^. When the cause of the event was undetermined, we verified if the event met criteria for embolic stroke of undetermined source (ESUS) definition^20^. The follow-up case-report-form collected details on the follow-up visit (i.e. date, remote or in presence) compliance to treatments, adverse events, effectiveness and safety outcome events, and mRS. For severity of the bleeding events, we referred to the definition of the Global Utilization of Streptokinase and Tissue Plasminogen Activator for Occluded Coronary Arteries (GUSTO) trial^21^.

Data were entered in an electronic anonymized database created with the Research Electronic Data Capture (REDCap) software^22,23^ hosted at University of L’Aquila. The study staff of the leading center remotely monitored progress and ensured integrity of data collection and completeness of follow-up through regular data quality checks as previously reported^13^. The investigators of the leading center – University of L’Aquila – had full access to all the data and take responsibility for its integrity and data analysis.

### 2.3 Outcomes

The primary effectiveness outcome was a composite of new stroke events (ischemic or hemorrhagic) or death due to vascular causes at 90 days. The primary safety outcome was a moderate -to-severe bleeding at 90 days.

Secondary outcomes were ischemic stroke (i.e. ischemic stroke or TIA), intracerebral hemorrhage (ICH), subarachnoid hemorrhage, other intracranial hemorrhage (i.e. subdural hematoma, epidural hematoma, or other), hemorrhagic infarction, myocardial infarction, vascular death, non-vascular death, any hospitalization, disability measured by the mRS, cause of DAPT discontinuation (i.e. adverse events, lack of compliance, other), severe bleeding, moderate bleeding, and mild bleeding. Secondary composite outcomes were any bleeding (i.e any of severe, moderate, or mild bleeding) and any death (i.e. any of death due to vascular or non-vascular causes).

Each local investigator adjudicated outcome events on review of medical charts. We reported outcomes in the entire study cohort and in pre-specified subgroups: age > or ≤ 65 years, body mass index (BMI) ≥ or <30, symptoms ≥ or <24 hours, presence of acute lesions at neuroimaging, NIHSS score ≤ or >3, ABCD^2^ score ≤ or >4, and loading dose.

### 2.4 Statistical analysis

The enrollment period was set at a minimum of 2 years with possibility to extend the duration if the minimal sample size was not reached. Using a 95% confidence interval, we estimated sample size of 1067 subjects to detect a conservative 50% proportion of primary effectiveness outcome with a two-sided 2.5% margin of error.

All analyses were performed according to the intention-to-treat principle in patients who completed the follow-up or had an outcome event within 90 days. We excluded patients who had an end of study visit prior than 80 days, those who underwent stenting procedures during the follow-up, and those who discontinued DAPT due to the diagnosis of atrial fibrillation or any other condition requiring anticoagulation – we excluded the latter group to avoid bias attributable to therapeutic switch. In case patients had multiple events, only the first outcome was used in the model.

We reported descriptive statistics about demographics, characteristics of the index event, and outcome events. Categorical data were reported as number and percentage. Continuous data had non-normal distribution at Kolmogorov-Smirnov test and were reported as median and interquartile range (IQR). Categorical and continuous data were compared across subgroups through the χ^2^ and Mann-Whitney U tests respectively.

To assess time distribution of outcome events during the follow-up, we performed survival analysis of primary effectiveness outcome and any bleeding. We further compared time to primary effectiveness outcome and any bleeding among patients who started DAPT within 12, 12-24 and after 24 hours from symptom onset with log-rank test.

Statistics were performed through SPSS version 20 (IBM Corp, Armonk, NY, USA) and R 3.3.0+; graphs were performed through GraphPad version 10.

## 3. Results

Out of 2278 patients enrolled between February 2021 and February 2023, 6.5% were lost to follow-up, 6.2% had a follow-up earlier than 80 days from the index event, 1.6% discontinued DAPT because of a diagnosis of atrial fibrillation or any other condition requiring anticoagulation, and 1.4% underwent stenting procedures. Therefore, we included in the analysis 1920 (84.3%) patients (Figure 1).

**Figure 1:**
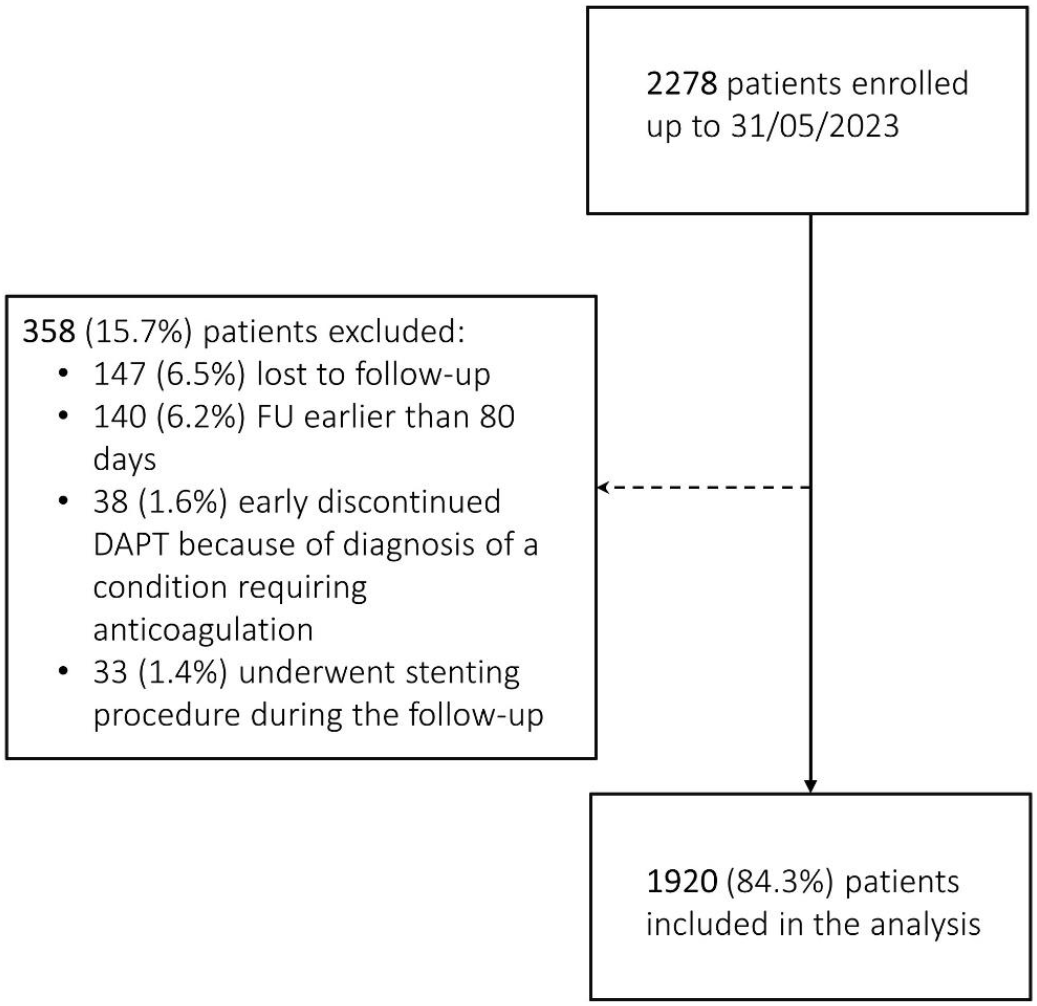
Consolidated Standards of Reporting Trials Diagram of Enrollment in the Study. Abbreviations: dual antiplatelet therapy (DAPT), follow-up (FU)

### 3.1 Demographics and characteristics of the index event

Most patients were male (65.4%) and Caucasian (97.8%), and median age was 72 years (interquartile range [IQR 62-79]). About one third of patients were on treatment with aspirin prior to the index event (33.8%), while fewer patients were on clopidogrel (6.9%) or ticagrelor (0.2%). Vascular risk factors were highly prevalent in our cohort with arterial hypertension being the most common (79.5%). Notably, 19.1% of patients had a history of prior ischemic event and 0.8% of prior ICH (Table 1a).

**Table 1a:** Baseline characteristics of study population.

| Characteristics | Cases with available data, N (%) | Overall study cohort |
| --- | --- | --- |
| Age, years, median (IQR) | 1920 (100) | 72 (62-79) |
| Female gender, N (%) | 1920 (100) | 665 (34.6) |
| Race, N (%) | 1920 (100) |  |
| Caucasian |  | 1877 (97.8) |
| Black |  | 10 (0.5) |
| Asian |  | 19 (1.0) |
| Other |  | 14 (0.7) |
| Blood pressure | 1739 (90.5) |  |
| Systolic |  | 150 (140-170) |
| Diastolic |  | 84 (75-90) |
| BMI, median (IQR) | 1920 (100) | 26 (24-28) |
| Current smoker, N (%) | 1920 (100) | 492 (25.6) |
| Hypertension, N (%) | 1920 (100) | 1527 (79.5) |
| Diabetes mellitus, N (%) | 1920 (100) | 521 (27.1) |
| Dyslipidemia, N (%) | 1920 (100) | 1160 (60.4) |
| Hypertriglyceridemia | 1920 (100) | 398 (20.7) |
| Cancer, N (%) | 1920 (100) | 234 (12.2) |
| Past* |  | 170 (8.9) |
| Current |  | 64 (3.3) |
| Previous ischemic event, N (%) | 1920 (100) | 366 (19.1) |
| TIA |  | 208 (10.8) |
| Ischemic stroke |  | 158 (8.3) |
| Previous intracerebral hemorrhage, N (%) | 1920 (100) | 15 (0.8) |
| Lobar |  | 3 (0.2) |
| Non-lobar |  | 8 (0.4) |
| Uncertain |  | 4 (0.2) |
| Myocardial infarction, N (%) | 1920 (100) | 183 (9.5) |
| Angina, N (%) | 1920 (100) | 62 (3.2) |
| Congestive heart failure, N (%) | 1920 (100) | 58 (3.0) |
| Peripheral Chronic Obliterative Arteriopathy, N (%) | 1920 (100) | 110 (5.7) |
| Use of antiplatelet prior to the index event, N (%) | 1920 (100) |  |
| Aspirin |  | 648 (33.8) |
| Clopidogrel |  | 132 (6.9) |
| Ticagrelor |  | 3 (0.2) |

Abbreviations: Body mass index (BMI); Interquartile range (IQR); Number (N); transient ischemic attack (TIA); \* absence of disease in the past 5 years

Most of patients received DAPT after an ischemic stroke (69.9%) with a median NIHSS score of 3 (IQR 2-4, range 0-28); 35.4% of patients had NIHSS score >3 and 11.8% NIHSS score >5 (Figure S1). A total of 30.1% of patients had a TIA with a median ABCD^2^ score of 5 (4-5); 20.1% had an ABCD^2^ score<4 (Figure S2) and 65.7% an ABCD^2^ score<6 without any symptomatic intracranial/extracranial stenosis. Eighteen point four percent of patients underwent a revascularization procedure, 1.0% had bridging therapy - intravenous thrombolysis (IVT) and endovascular thrombectomy (EVT) - and 0.3% of patients had endarterectomy and either IVT or EVT. In total, 16.2% of patients received IVT, 1.9% EVT, and 2.4% endarterectomy (Table 1b).

**Table 1b:**
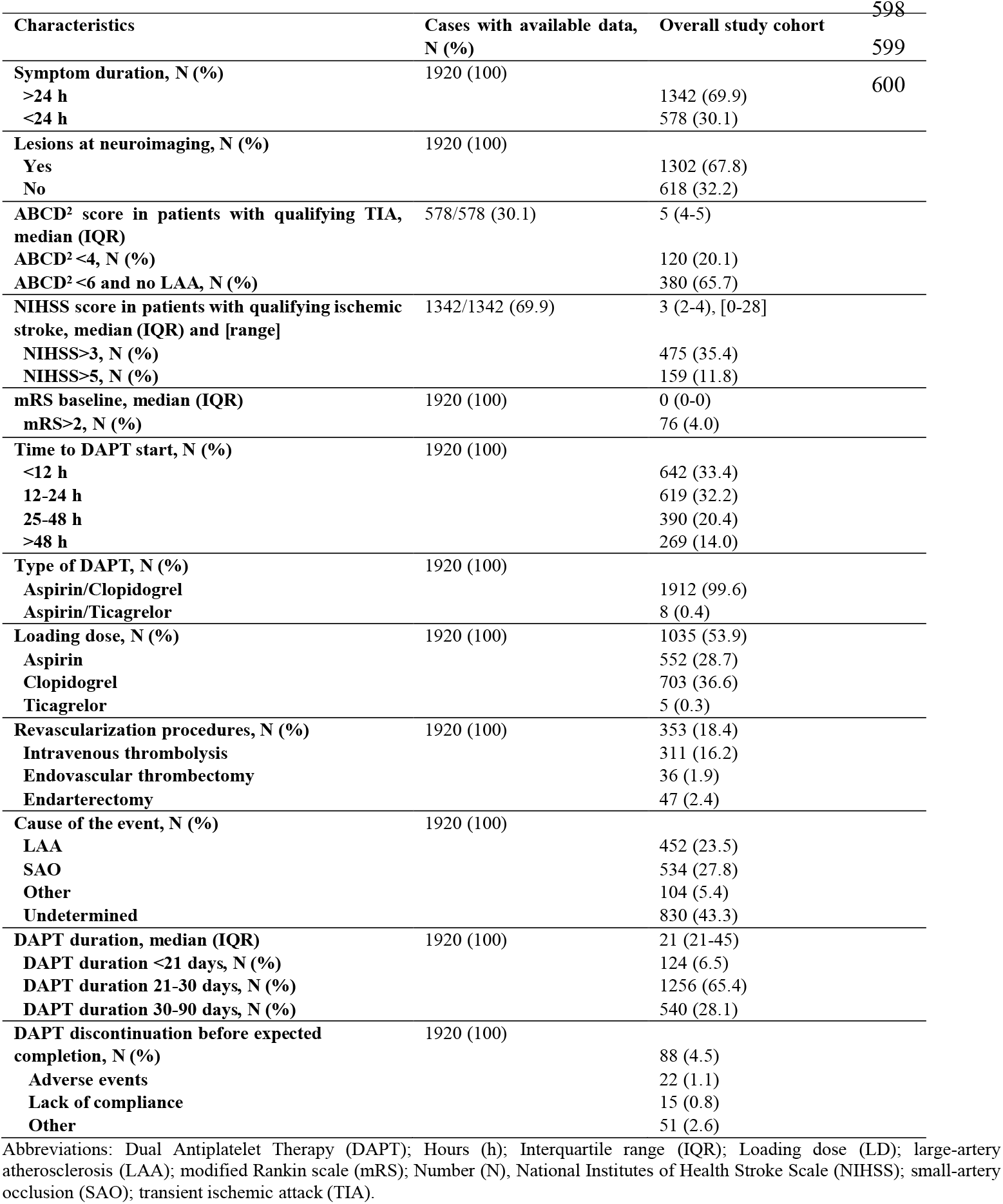
Characteristics the index event.

The cause of the event was in most of the cases undetermined (43.3%) followed by small-artery occlusion (SAO) (27.8%), large-artery atherosclerosis (LAA) (23.5%) and other determined causes (5.4%) (Table 1b). Among the other determined causes, dissection accounted for 27.9% of cases, vasculitis for 3.8%, hypercoagulability for 4.8%, and other mixed causes for the remaining 63.5% of cases. Among the undetermined events, 32.5% were classified as ESUS and in 11% of cases physicians detected a patent foramen ovale (PFO).

### 3.2 DAPT regimen

The most prescribed DAPT was clopidogrel/aspirin (99.6%), with majority of patients starting the therapy within 12 hours from symptom onset (33.4%) and 14.0% after 48 hours (Table 1b).

A total of 53.9% of patients received a loading dose of any antiplatelets with clopidogrel being the most common (36.6%). DAPT median duration was 21 days (21-45); 4.5% of patients discontinued DAPT before the expected completion date: in 1.1% of cases because of adverse events and in 0.7% due to lack of compliance. Bleedings were the most common adverse events leading to treatment discontinuation (Table 1b).

Aspirin was the most frequent antiplatelet prescribed after DAPT cessation (62.9%) followed by clopidogrel (35.0%), while 2.1% of patients discontinued all the antiplatelets. Out of 648 patients treated with aspirin prior to the index event, 62.9% received clopidogrel after DAPT cessation, while 62.1% of the 132 patients treated with clopidogrel prior to the index event received aspirin after DAPT cessation.

### 3.3 Effectiveness and safety outcomes

During the 90-day follow-up, a primary effectiveness outcome occurred in 3.9% of patients and a primary safety outcome occurred in 0.6% of patients; 3.3% of patients had an ischemic recurrence (2.0% had an ischemic stroke and 1.3% a TIA) and 0.2% had an ICH. In addition, 0.2% of patients had a subarachnoid hemorrhage and 0.1% other intracranial bleedings. Myocardial infarction occurred in 0.2% of patients and 3.3% required a new hospitalization. A total of 78.8% of patients had mRS of 0-1 and median mRS was 0 (IQR 0-1) at 90 days (Table 2).

**Table 2:** Primary and secondary effectiveness and safety outcome in READAPT, CHANCE, POINT and THALES.

| Outcome | READAPT* | CHANCE<sup>6</sup> | POINT<sup>7</sup> | THALES<sup>8</sup> |
| --- | --- | --- | --- | --- |
| Sample size | 1920 | 2584 | 2432 | 5493 |
| *Primary outcomes* |  |  |  |  |
| Primary effectiveness outcome↓, N (%) | 74 (3.9) | 212 (8.2) | 121 (5.0) | 303 (5.5) |
| Primary safety outcome$\S$ N (%) | 12 (0.6) | 7 (0.3) | 23 (0.9) | 28 (0.5) |
| *Key secondary outcomes* |  |  |  |  |
| Ischemic event, N (%) | 64 (3.3) | 243 (9.4) | 112 (4.6) | 276 (5.0) |
| Ischemic stroke | 38 (2.0) | 204 (7.9) | 112 (4.6) | 276 (5.0) |
| TIA | 26 (1.3) | 39 (1.5) | NA | NA |
| Hemorrhagic transformation, N (%) | 19 (1.0) | NA | NA | NA |
| Symptomatic | 2 (0.1) |  |  |  |
| Asymptomatic | 17 (0.9) |  |  |  |
| Intracranial hemorrhage, N (%) | 3 (0.2) | 8 (0.3) | 5 (0.2) | 22 (0.4) |
| Subarachnoid hemorrhage, N (%) | 2 (0.1) | NA | NA | NA |
| Other intracranial hemorrhage, N (%) | 2 (0.1) | NA | NA | NA |
| Myocardial infarction, N (%) | 3 (0.2) | 3 (0.1) | 10 (0.4) | NA |
| Death, N (%) | 8 (0.5) | 10 (0.4) | 18 (0.7) | 36 (0.7) |
| Vascular | 3 (0.2) | 6 (0.2) | 6 (0.2) |  |
| Non-vascular | 5 (0.3) | 4 (0.2) | 12 (0.5) |  |
| Severe bleeding, N (%) | 9 (0.5) | 4 (0.2) | 17 (0.7) | 28 (0.5) |
| Moderate bleeding, N (%) | 3 (0.2) | 3 (0.1) | NA | 36 (0.7%)<sup>a</sup> |
| Mild bleeding, N (%) | 40 (2.1) | 30 (1.2) | 40 (1.6) | 36 (0.7%)<sup>a</sup> |
| Any bleeding, N (%) | 52 (2.7) | 60 (2.3) | NA | NA |
| Hospitalization, N (%) | 63 (3.3) | NA | NA | NA |
| mRS, median (IQR) | 0 (0-1) | NA | NA | NA |

Abbreviations: modified Rankin scale (mRS); Number (N), transient ischemic attack (TIA) ↓ primary efficacy outcome in CHANCE was new stroke event (ischemic or hemorrhagic), in POINT was the composite of ischemic stroke, myocardial infarction, or death from ischemic vascular causes, and in THALES was the composite of stroke (ischemic stroke, hemorrhagic stroke) or death; $\S$ primary safety outcome in CHANCE was Moderate-severe bleeding event, in POINT was major hemorrhage, and in THALES; <sup>a</sup>moderate or mild bleeding; \*all patients were evaluable for outcome analysis

Overall, 2.7% of patients had any bleeding event. Severe bleedings occurred in 0.5% of patients, moderate in 0.2%, and mild bleedings in 2.1%. Death occurred in 0.5% of patients, 0.2% due to vascular causes and 0.3% due to non-vascular causes (Table 2). Hemorrhagic infarction occurred in 0.9% of patients and was symptomatic only in 0.1% of cases.

Most of the outcomes occurred while patients were still on DAPT treatment: 60.8% of primary effectiveness outcomes, 58.3% of primary safety outcomes, and 61.5% of any bleedings. Notably, 2.0% of patients had multiple outcome events; additional details are available in the supplemental material (Table S1).

### 3.4 Effectiveness and safety in subgroups of interest

Primary effectiveness outcome was more frequent among those without acute lesion at neuroimaging (5.5%) compared with those with lesions at neuroimaging (3.1% P=0.010) and among patients with NIHSS score ≤5 (4.0%) compared with NIHSS score >5 (0.6%, P=0.033) (Figure 2A). A primary safety outcome occurred more often in patients with NIHSS score >3 and compared with NIHSS≤3 (P=0.019). Any bleeding was more common among patients >65 years (3.2%) compared with those ≤65 years (1.7%, P=0.055) (Table S3) (Figure 2B). We found no statistically significant difference in the other subgroups defined according to BMI, symptom duration, and loading dose.

**Figure 2:**
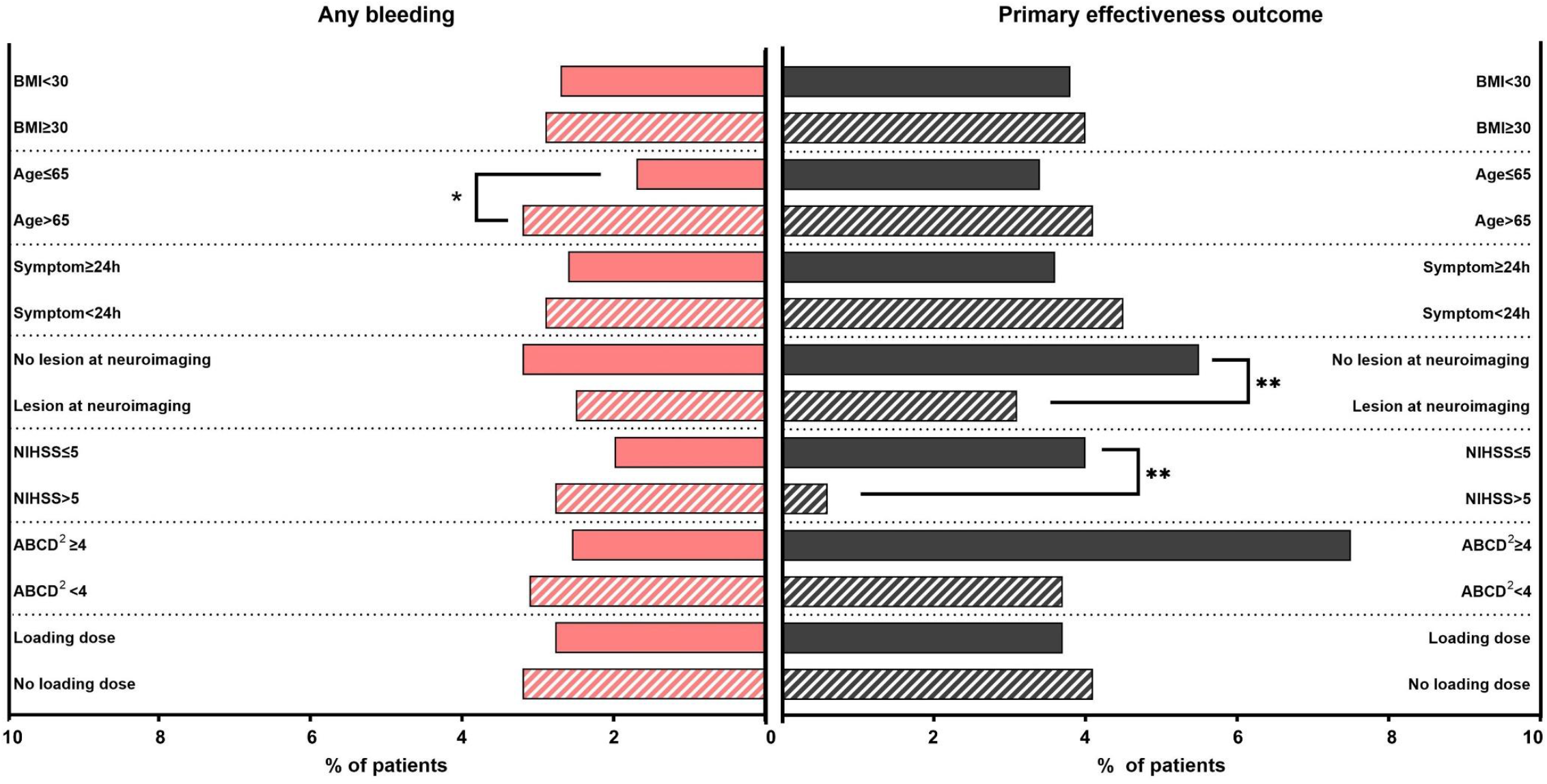
rates of any bleeding and primary effectiveness outcomes across pre-specified subgroups. Abbreviations: Body mass index (BMI), Hours (h); National Institutes of Health Stroke Scale (NIHSS).

### 3.5 Cumulative probabilities of primary effectiveness outcome and any bleeding according to time to DAPT start

Out of the 74 primary effectiveness outcomes, 18.9% occurred the day after the index event and 28.3% within 7 days; lower rates occurred in the following weeks until day 90 (Figure 3A). There were no differences in the occurrence of primary effectiveness outcome among patients who started DAPT within 12 hours, between 12 and 24 hours, and after 24 hours from symptom onset (P=0.3) (Figure 3B).

**Figure 3:**
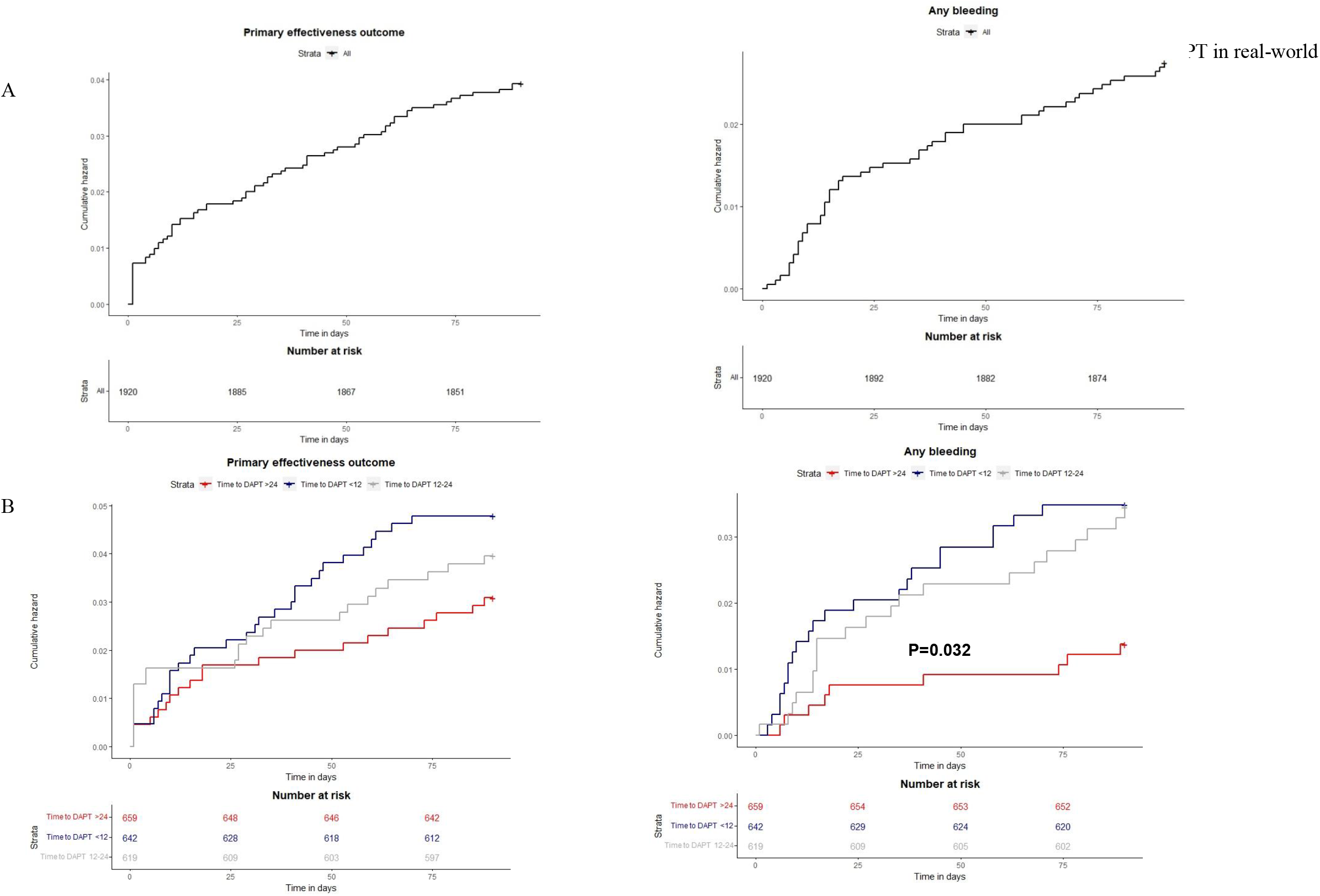
Cumulative Hazards of the primary effectiveness outcome and any bleeding in the overall study cohort (A) and in subgroups of patients defined according to time to dual antiplatelet therapy start (B).

Out of the 52 any bleeding events, 15.4% occurred within the first week from index event, 23.1% in the second week, and 11.5% in the third week (Figure 3A). Number and proportion of any bleeding gradually decreased in the following weeks. Patients treated with DAPT within 12 hours and between 12-24 hours were at higher risk of any bleeding than those who started DAPT after 24 hours from symptom onset (P=0.032) (Figure 3B).

## 4. Discussion

READAPT study provided data on effectiveness and safety of short-term DAPT in secondary prevention of non-cardioembolic minor ischemic stroke or high-risk TIA in real-world. As previously reported, READAPT cohort included patients who did not meet RCTs definitions of “minor” ischemic stroke or “high-risk” TIA, namely NIHSS or ABCD^2^ scores were above or below the cut-off values^13^. Additionally, some patients received DAPT after a revascularization procedure, did not receive a loading dose or had a later initiation of DAPT. Despite the broader use of DAPT in our cohort, we did not identify any safety concern. Effectiveness outcomes turned out to be even lower than anticipated on RCTs results.

During the 90-day follow-up, we observed 3.9% primary effectiveness outcomes – a recurrent ischemic event, vascular death, or an intracranial bleeding – mainly driven by ischemic recurrences which occurred in 3.3% of patients^6-8^. Most of the recurrences occurred early after the index event thus reinforcing the need to start DAPT as early as possible in eligible patients. We found a lower rate of 90-day ischemic recurrences in READAPT than in other real-world studies (3.8-4.7%), which included both patients treated with DAPT and with other preventative therapies^3,5^. The same rate was also lower than those reported in RCTs^6-8^ (Table 2) for several possible reasons. For instance, investigators might have missed recurrences presenting with mild or worsening of preexisting symptoms especially after patient discharge. In addition, some early recurrences might have occurred before DAPT start despite patients who started DAPT within 12, between 12-24 and after 24 hours from symptom onset had similar risks of primary effectiveness outcomes. The effectiveness of DAPT might have been greater than in RCTs due to patients’ characteristics and concomitant therapies. In READAPT, Caucasian ethnicity was the most represented, rates of Asian and Black patients were smaller than those of POINT trial^7^, which was the RCT on DAPT with the greatest multiethnic representation^7^. A greater effectiveness of clopidogrel-DAPT - the most used type of DAPT in our cohort - might be due to the scarce representation of Asian patients, who more frequently are carriers of loss of function CYP2C9 polymorphisms^24,25^, and of Black patients, who proved at higher risk of ischemic recurrences^26^. Intensive blood pressure lowering agents and/or statins according to patients’ comorbidities might have boosted DAPT effectiveness in our cohort. Lastly, READAPT patients might have presented a higher vascular risk profile than those in RCTs according to median age, and prevalence of arterial hypertension and prior vascular events. Previous real-world studies comparing subgroups of patients according to their risk profile have shown that DAPT was more effective in those at higher compared with those at lower risk ^27,28^.

DAPT had a similar safety profile in READAPT compared to RCTs^6-8^ (Table 2). We observed proportion of primary safety events – moderate to severe bleeding – similar to those reported in the trials, but mild bleedings were about one percentage point higher^6-8^. Furthermore, bleedings were among the most frequent adverse events leading to treatment discontinuation. Time-to-event analysis showed that DAPT start within 12 hours from symptom onset was associated with a higher risk of any bleeding possibly due to a higher rate of hemorrhagic transformation or intracranial bleedings in these patients. A proper evaluation of patients’ hemorrhagic risks including an assessment of the volume of ischemic lesions might minimize hemorrhagic complications of DAPT. The exact window for DAPT start after an ischemic event is yet to be defined, INSPIRES and a sub-analysis of POINT showed that DAPT was still beneficial if administered within 72 hours from symptom onset^9,29^, thus a delayed treatment start might be considered in specific cases. Mortality was not a concern in our cohort, and the rate of vascular death was comparable to RCTs^6-8^.

READAPT study showed that the real-world use of DAPT is broader than in RCTs. Indeed, we included patients whose DAPT benefit/risk profile is unclear. For instance, those treated with revascularization procedures and those outside the RCTs boundaries of minor stroke or high-risk TIA, which, in clinical practice, are perceived at high risk for ischemic events and at a low risk of bleeding. Indeed, about 35% of patients had an NIHSS score >3, 20% an ABCD^2^ score<4, and 18% received a revascularization procedure. We found no significant difference in primary effectiveness outcome among those with ABCD^2^ score ≥ or < 4 supporting the need for a comprehensive vascular risk evaluation not only based on ABCD^2^ score^30^. Conversely, patients with NIHSS score >5 had a lower rate of primary effectiveness outcome compared with those with NIHSS score ≤5 in line with a previous real-world study which identified NIHSS 3-10 as a predictor of DAPT effectiveness^31^. The lower number of patients with NIHSS score >5 requires cautious interpretation of this result. In addition, the possible greater DAPT effectiveness among patients with more severe stroke is offset by the increased rate of primary safety outcome we observed in patients with NIHSS score >3. Regarding safety, patients older than 65 years had a higher rate of any bleeding events. Concomitant comorbidities such as hypertension, renal or liver dysfunctions, which are more frequent among the elderly, might increase the risk of systemic bleeding. Similarly, a metanalysis showed an increased risk of severe bleeding among patients ≥ 65 years treated with DAPT compared with SAPT^32^.

However, the metanalysis mainly included data of RCTs that evaluated long-term DAPT efficacy with various combinations of antiplatelets and dosages^32^. The safety of short-term DAPT in the elderly is yet to be investigated in controlled trials: it is unclear whether the bleeding risk linearly increases with the age, as shown for long-term SAPT with aspirin^33^, and whether other cofactors might play a role.

In terms of DAPT benefit/risk timeseries, we found that most of the primary effectiveness outcomes occurred within the first week, while the rates of any bleeding peaked during the second week and then remained constant up to day 90. A time-course analysis of CHANCE showed that DAPT reduced the 1-week risk of new ischemic stroke by approximately 35% and numerically increased the risk of any bleeding during the first 4 weeks^34^. Both in READAPT and CHANCE subgroup analyses there was an increase of hemorrhagic risk during the second week, but we did not observe a similar continuous increasing rate of any bleeding over time. This different trend in READAPT might be due to the low number of any bleeding events and/or lack of treatment compliance.

READAPT adopted rigorous procedures to ensure accuracy, completeness, and quality of data such as the use of specific electronic case-report-forms and regular quality checks. Among the limitations of the study, the geographical setting restricted to Italy and the mainly Caucasian population did not allow to generalize our results to other countries. In addition, our results might not be an exact representation of the whole national population as recruitment was not equal across all participating centers with larger contributions of centers located in urban areas. The observational design of the study might have led to an underestimation of the outcomes. The study was underpowered to identify predictors of outcome events, thus, we performed only descriptive statistics. Physicians reported the outcomes and the neuroimaging characteristics based on patients’ record that we did not have had direct access to. Therefore, the appropriateness of outcome adjudication was upon local investigators. Lastly, we excluded from the study patients with minor stroke due to cardioembolic sources with a possible selection bias. We further excluded those who discontinued DAPT because of a diagnosis of atrial fibrillation as the scope of the study was to evaluate the effectiveness of DAPT in patients with non-cardioembolic causes. However, we cannot exclude that an occult cardioembolism might have caused some of the index or recurrent ischemic event.

## 5. Conclusions

Use of short-term DAPT for secondary prevention of ischemic stroke or TIA is broader in real-world than in clinical trials as physicians prescribed the treatment to patients outside the cut-off values for minor stroke or high-risk TIA and/or undergoing revascularization procedures. Also, time-to-DAPT start from symptom onset and use of loading doses differ from clinical trials. READAPT study showed that, in real-world setting, there are no emerging concerns and DAPT maintains safe even in a broader population and following slightly different procedures. Indeed, treatment maintains benefits in patients with ischemic stroke of moderate severity, low-risk TIA, and in those who started the treatment after 24 hours from symptom onset; only elderly appear at higher risk of bleeding. Additional clinical trials are required to evaluate the benefits of short-term DAPT in this broader population. Based on READAPT results, we suggest considering DAPT in patients deemed at high risk even if not strictly meeting criteria for minor stroke or high-risk TIA and promptly starting it after symptom onset, since most of the outcome events occurred early after the index event.

## Data Availability

Data are available upon reasonable request to the Corresponding Author

## List of abbreviations and acronyms

ABCD^2^: Age, Blood pressure, Clinical features, Duration of TIA, presence of Diabetes
CHANCE: Clopidogrel in High-Risk Patients With Acute Non Disabling Cerebrovascular Events trial
DAPT: dual antiplatelet therapy
EVT: endovascular thrombectomy
GUSTO: Global Utilization of Streptokinase and Tissue Plasminogen Activator for Occluded Coronary Arteries
ICH: intracerebral hemorrhage
ISA-AII: Italian Stroke Association-Associazione Italiana Ictus
IVT: intravenous thrombolysis
LAA: large-artery atherosclerosis
mRS: modified Rankin Scale
NIHSS: National Institutes of Health Stroke Scale
POINT: Platelet Oriented Inhibition in New TIA and Minor Ischemic Stroke trial
READAPT: REAl-life study on short-term Dual Antiplatelet treatment in Patients with ischemic stroke or Transient ischemic attack study
RCT: Randomized controlled trials
SAO: small-artery occlusion
THALES: The Acute Stroke or Transient Ischemic Attack Treated with Ticagrelor and Acetylsalicylic acid for Prevention of Stroke and Death trial
TIA: transient ischemic attack
TOAST: Trial of Org 10172 in acute stroke treatment
WHO: World Health Organization

## 8. Acknowledgements

The Authors wish to thank all the study patients for their kind cooperation. Dr Sacco conceived the study and its design; Drs Ricci and Toni provided critical revision to the study protocol; Drs De Matteis coordinated data collection, performed the acquisition, analysis, and interpretation of data; Drs Ornello, Foschi and Romoli provided support in statistical analysis; Drs Sacco, De Matteis, and Ornello drafted the manuscript; Drs De Matteis, De Santis, Foschi, Romoli, Tassinari, Saia, Cenciarelli, Bedetti, Padiglioni, Censori, Puglisi, Vinciguerra, Guarino, Barone, Zedde, Grisendi, Diomedi, Bagnato, Petruzzellis, Mezzapesa, Di Viesti, Inchingolo, Cappellari, Zenorini, Candelaresi, Andreone, Rinaldi, Bavaro, Cavallini, Morau, Querzani, Terruso, Mannino, Pezzini, Frisullo, Muscia, Paciaroni, Mosconi, Zini, Palmieri, Cupini, Marcon, Tassi, Sanzaro, Paci, Viticchi, Orsucci, Falcou, Diamanti, Tarletti, Nencini, Rota, Sepe, Ferrandi, Caputi, Volpi, La Spada, Beccia, Rinaldi, Mastrangelo, Di Blasio, Invernizzi, Pelliccioni, De Angelis, Bonanni, Ruzza, Caggia, Russo, Tonon, Acciarri, Anticoli, Roberti, Manobianca, Scaglione, Pistoia, Fortini, De Boni, Sanna, Chiti, Barbarini, Caggiula Masato, Del Sette, Passarelli, Bongioanni, Toni, Ricci, Sacco acquired data, provided critical revision in the analysis and interpretation of data, approved the final manuscript, and agreed to be accountable for all aspects of the work in ensuring that questions related to the accuracy or integrity of any part of the work are appropriately investigated and resolved.

## 9. Source of funding

READAPT is a non-profit study.

## 10. Disclosures

Dr Zini reports compensation from Angels Initiative, Boehringer-Ingelheim, Daiichi Sankyo for consultant services; from Angels Initiative, Boehringer-Ingelheim, CSL Behring for speaking honoraria or other education services; from Daiichi Sankyo for meeting; from Bayer, and Astra Zeneca for participation on a Data Safety, Monitoring Board or Advisory Board; and he is member of ESO guidelines, ISA-AII guidelines, and IRETAS steering committee. Dr Ornello reports grants from Novartis and Allergan; compensation from Teva Pharmaceutical Industries, Eli Lilly and Company, and Novartis for other services; and travel support from Teva Pharmaceutical Industries. Dr Sacco reports compensation from Novartis, NovoNordisk, Allergan, AstraZeneca, Pfizer Canada, Inc, Eli Lilly and Company, Teva Pharmaceutical Industries, H. Lundbeck A/S, and Abbott Canada for consultant services; employment by Università degli Studi dell’Aquila; and compensation from Novartis for other services. Dr Paciaroni reports compensation from Daiichi Sankyo Company, Bristol Myers Squibb, Bayer, and Pfizer Canada, Inc, for consultant services. Dr Toni reports compensation from Alexion, AstraZeneca, Medtronic, and Pfizer for consultant services and participation on a Data Safety, Monitoring Board or Advisory Board. The other authors report no conflicts.

## 11. Ethics statement

The study was approved by the Internal Review Board of the University of L’Aquila (Italy) and patients gave written informed consent according to the Declaration of Helsinki.

## 12. List of supplemental material

Figure S1

Figure S2

Figure S3

Table S1

Table S2

Table S3

READAPT group authorship

## Notes

** A list of all authors in the READAPT Study Group is in the Supplemental Material.

### Competing Interest Statement

The authors have declared no competing interest.

### Funding Statement

The study is a non-profit study.

### Author Declarations

Internal Review Board of the University of L'Aquila (Italy) has cleared the study with the number 03/2021

